# Prediabetes as a risk factor for all-cause and cause-specific mortality: a prospective analysis of 115,919 adults without diabetes in Mexico City

**DOI:** 10.1101/2024.04.15.24305840

**Authors:** Carlos A. Fermín-Martínez, Omar Yaxmehen Bello-Chavolla, César Daniel Paz-Cabrera, Daniel Ramírez-García, Jerónimo Perezalonso-Espinosa, Luisa Fernández-Chirino, Arsenio Vargas-Vázquez, Juan Pablo Díaz-Sánchez, Padme Nailea Méndez-Labra, Alejandra Núñez-Luna, Martín Roberto Basile-Alvarez, Paulina Sánchez-Castro, Fiona Bragg, Louisa Gnatiuc Friedrichs, Diego Aguilar-Ramírez, Jonathan R. Emberson, Jaime Berumen-Campos, Pablo Kuri-Morales, Roberto Tapia-Conyer, Jesus Alegre-Díaz, Jacqueline A. Seiglie, Neftali Eduardo Antonio-Villa

## Abstract

**BACKGROUND:** Prediabetes has been associated with increased all-cause and cardiovascular mortality. However, no large-scale studies have been conducted in Mexico or Latin America examining these associations.

**METHODS:** We analyzed data from 115,919 adults without diabetes (diagnosed or undiagnosed) aged 35-84 years who participated in the Mexico City Prospective Study between 1998 and 2004. Participants were followed until January 1^st^, 2021 for cause-specific mortality. We defined prediabetes according to the American Diabetes Association (ADA, HbA_1c_ 5.7% to 6.4%) and the International Expert Committee (IEC, HbA_1c_ 6.0-6.4%) definitions. Cox regression adjusted for confounders was used to estimate all-cause and cause-specific mortality rate ratios (RR) at ages 35-74 years associated with prediabetes.

**FINDINGS:** During 2,085,392 person-years of follow-up (median in survivors 19 years), there were 6,810 deaths at ages 35-74, including 1,742 from cardiovascular disease, 892 from renal disease and 108 from acute diabetic crises. Of 110,405 participants aged 35-74 years at recruitment, 28,852 (26%) had ADA-defined prediabetes and 7,203 (7%) had IEC-defined prediabetes. Compared with those without prediabetes, individuals with prediabetes had higher risk of all-cause mortality at ages 35-74 years (RR 1.13, 95% CI 1.07-1.19 for ADA-defined prediabetes and RR 1.28, 1.18-1.39 for IEC-defined prediabetes), as well as increased risk of cardiovascular mortality (RR 1.22 [1.10-1.35] and 1.42 [1.22-1.65], respectively), renal mortality (RR 1.35 [1.08-1.68] and 1.69 [1.24-2.31], respectively), and death from an acute diabetic crisis (RR 2.63 [1.76-3.94] and 3.43 [2.09-5.62], respectively). RRs were larger at younger than at older ages, and similar for men compared to women. The absolute excess risk associated with ADA and IEC-defined prediabetes at ages 35-74 accounted for6% and 3% of cardiovascular deaths respectively, 10% and 5% of renal deaths respectively, and 31% and 14% of acute diabetic deaths respectively.

**INTERPRETATION:** Prediabetes is a significant risk factor for all-cause, cardiovascular, renal, and acute diabetic deaths in Mexican adults. Identification and timely management of individuals with prediabetes for targeted risk reduction could contribute to reducing premature mortality from cardiometabolic causes in this population.

**FUNDING:** Wellcome Trust, the Mexican Health Ministry, the National Council of Science and Technology for Mexico, Cancer Research UK, British Heart Foundation, UK Medical Research Council. Instituto Nacional de Geriatría (Mexico City).

**RESEARCH IN CONTEXT:** *Evidence before this study:* We conducted a literature search in PubMed to identify articles published in English before February 27^th^, 2024 that reported on prospective studies examining the association between prediabetes with all-cause or cause-specific mortality or progression to diabetes in a Mexican or Latin American population, using the terms (“prediabetes” OR “impaired fasting glucose” OR “impaired glucose tolerance”) AND (“mortality” OR “death”) AND (“Mexico” OR “Mexican” OR “Latin America” OR “Latin American”). There were no studies examining risk associated with prediabetes definitions and mortality among adults in Mexico. We identified one study from Peru that included 988 participants and investigated only all-cause mortality for impaired fasting glucose and HbA_1c_-based definitions of prediabetes from ADA and IEC; this study reported increased mortality risk related to ADA-defined prediabetes based on HbA_1c_ measures. Generalizability of these findings to other Latin American countries and regions with distinct cardiometabolic profiles in unclear.

*Added value of this study:* Our study included 115,919 participants without diabetes from Mexico City, of whom (26%) had ADA-defined prediabetes and 7,203 (7%) had IEC-defined prediabetes. We found that prediabetes is associated with higher risks of all-cause and cause-specific mortality (cardiovascular, renal, and acute diabetic causes) than among participants without prediabetes. We found RRs to be larger at younger than at older ages, and largely similar for men compared to women. Among those without previously diagnosed diabetes, we found that the excess risk associated with ADA- and IEC-defined prediabetes at ages 35-74 years accounted for 6% and 3% of cardiovascular deaths, 10% and 5% of renal deaths, and 31% and 14% of acute diabetic deaths, respectively. .

*Implications of all the available evidence:* Our results show that prediabetes is a significant risk factor for cardiovascular, kidney, and acute diabetic deaths among Mexican adults and accounts for a notable fraction of such deaths. Identification of individuals with prediabetes should be prioritized for optimized management to improve cardiometabolic outcomes in Mexican adults.

## INTRODUCTION

Although the definition of prediabetes is the subject of continuous debate, it is well-recognized as a risk factor for development of type 2 diabetes and long-term adverse cardio-metabolic outcomes^1–3^. Prediabetes is an early stage indicator of glycemic dysregulation, and individuals with prediabetes often present with other traits of the metabolic syndrome (i.e. dysfunctional adipose tissue accumulation, dyslipidemia, insulin resistance)^4,5^. Despite its pathophysiological relevance and the overwhelming increase in the incidence of metabolic syndrome in susceptible populations, routine screening for prediabetes is disputed, in part due to a lack of consensus on how it should be defined^1^. Growing evidence supports that beyond its well-known association with progression to diabetes, prediabetes also increases the risk of cardiovascular disease and mortality^3^. Nevertheless, data on prediabetes-associated complications beyond progression to diabetes are limited, particularly in low- and middle-income countries (LMICs)^6^.

In Mexico, the prevalence of diabetes has increased at an alarming rate in recent decades^7^. This trend can be attributed to a complex interplay of underlying genetic risk and changes in environmental factors^8–10^, which have led to a steady increase in central obesity, impaired glucose tolerance and insulin resistance^11–14^. We recently described the trends in prediabetes prevalence in the Mexican population^7^; we found that although lower cut-offs of glycemic markers may be more adequate for screening strategies, they may not fully reflect the long-term health risk associated with prediabetes^15^. Notably, data on the risk conferred by prediabetes for all-cause and cause-specific mortality in Mexico are lacking. In this report, we aim to examine prediabetes as a risk factor for all-cause and cause-specific mortality in Mexican adults previously enrolled in the Mexico City Prospective Study (MCPS) and followed long-term for cause-specific mortality.

## METHODS

### Study design and participants

Details on recruitment and follow-up of participants in MCPS have been described previously^16^. Briefly, participants living in two municipalities in Mexico City (Coyoacán and Iztapalapa) aged ≥35 years were invited to participate in the study between 1998 and 2004. The study was approved by Ethics Committees at the Mexican Ministry of Health, the Mexican National Council for Science and Technology, and the University of Oxford, UK. All participants provided written informed consent.

### Data collection

Sociodemographic, health-related and lifestyle information were collected using an electronic questionnaire by trained nurses. All participants had their height, weight, hip circumference (HC), waist circumference (WC) and sitting blood pressure measured using calibrated instruments and standard protocols. A non-fasting venous blood sample was obtained and glycosylated hemoglobin (HbA_1c_) levels were measured using a validated high-performance liquid chromatography method^17^ on HA-8180 analyzers with calibrators traceable to International Federation of Clinical Chemistry standards^18^.

### Mortality follow-up

Participants were followed for cause-specific mortality through probabilistic linkage (based on name, age, and sex) to the Mexican electronic death registry. Death registration in Mexico City is reliable and complete, with nearly all deaths medically certified^19^. Diseases recorded on death certificates are coded using the International Statistical Classification of Diseases and Related Health Problems, Tenth Revision, with subsequent review by study clinicians (unaware of baseline information) to recode, where necessary, the underlying cause of death. Participant deaths for the present study were tracked up to January 1^st^, 2021. We grouped deaths as cardiac (mainly ischemic heart disease), cerebrovascular, other vascular (mainly peripheral arterial disease and thromboembolism), renal, acute diabetic crises (diabetic coma or ketoacidosis), hepatobiliary (mainly cirrhosis), other gastrointestinal (mainly peptic ulcer and gastrointestinal infections), neoplastic, respiratory (mainly pneumonia and chronic obstructive pulmonary disease), and external, ill-defined, or other causes of death. Full details on ICD-10 codes for individual causes of death are reported in **Supplementary Material**.

### Prediabetes and diabetes definitions

Prediabetes was defined according to the HbA1c cut-offs recommended by the American Diabetes Association (ADA) (HbA_1c_ 5.7-6.4%)^20^ and by the International Expert Committee (IEC, HbA_1_c 6.0-6.4%).^21^ We excluded individuals with previously diagnosed diabetes (self-reported medical diagnosis or use of glucose-lowering pharmacotherapy), individuals who reported regular use of glucose-lowering pharmacotherapy regardless of prior diagnosis, and individuals with undiagnosed diabetes (HbA1c ≥6.5% in someone without a prior diabetes diagnosis). To explore the mortality risks in the population with HbA1c levels between the ADA and IEC cutoff points, additional analyses categorized participants according to HbA_1_c levels <5.7%, 5.7-5.9%, and 6.0-6.4%.

### Statistical analysis

We limited our analysis to the population with prediabetes with complete data on HbA_1_c, mortality, and covariates. To limit the potential effects of reverse causation, we excluded those with self-reported chronic comorbidities at baseline (ischemic heart disease, stroke, chronic kidney disease, chronic obstructive pulmonary disease, cirrhosis, or cancer).^22^ We evaluated the prevalence of ADA and IEC-defined prediabetes in the remaining participants separately by age (in 5-year groups) and sex. Uniformly age- and sex-standardized death rates were estimated for those with and without ADA and IEC-defined prediabetes, as well as for the complementary HbA1c categories (above), and reported as events per 1,000 person-years.

The primary analyses were restricted to deaths at ages 35-74 years (i.e., deaths one might consider to be ‘premature’), but some analyses of deaths at ages 75-84 were also performed. Cox proportional hazards regression was used to estimate mortality rate ratios (RRs, estimated from the Cox hazard ratios) for all-cause and cause-specific mortality associated with prediabetes and HbA_1c_ levels. Cox models yield cause-specific log hazard ratios that provide a weighted average of the log mortality RRs in different time periods which, irrespective of the proportional hazards assumption, provide useful summary statistics of the average death RRs during the study. Participants who did not die from the cause under study were censored at the earliest of: death from an alternative cause; the end of the risk period under consideration (e.g., age 75 years for analyses of risk during ages 35-74 years); or the end of follow-up for mortality (January 1^st^ 2021).

All models were stratified by sex and age at risk (in 5-year groups) using the Lexis expansion (*Epi* R package)^23,24^ and were progressively adjusted for confounders in the following order: 1) municipality of residence (Coyocán or Iztapalapa) (region-level variations in population characteristics have been known to influence mortality related to cardiometabolic diseases in Mexico^25^), and 2) education level (university/college, or other), physical activity (none, regular), smoking (never, former, current), and alcohol consumption (never, former, current). Sensitivity analyses further adjusted for adiposity markers (body mass index [BMI] and waist-to-height ratio [WHtR]). Subgroup analyses estimated mortality RRs separately by age at risk and sex. The excess mortality attributable to prediabetes (in a population without previously diagnosed or undiagnosed diabetes) was estimated by applying the cause-specific mortality RRs to the number of deaths among those with prediabetes. All analyses were conducted using R (version 4.2.1).

## RESULTS

Of 159,517 participants who were originally recruited, 43,598 (27.3%) were excluded from the present analyses. These comprised 29,948 (18.8%) with diagnosed or undiagnosed diabetes at recruitment, a further 7,068 (4.4%) with missing data on HbA1c or covariates, or with uncertain mortality linkage, an additional 5,173 (3.2%) with chronic comorbidities at baseline, and1,409 (0.9%) aged ≥85 years at recruitment (**Supplementary** Figure 1). Of the remaining 115,919 participants, 110,405 (95.2%) were aged 35–74 years and 5,514 (4.8%) were aged 75–84 years at recruitment.

Participant characteristics at baseline, stratified by HbA1c categories, are summarized in Table 1. Participants in the highest HbA1c category (HbA1c 6.0%-6.4%) were older (mean age 54 years), predominantly female (70.%), had higher BMI (mean 32 kg/m^2^), and systolic blood pressure (mean 132 mmHg) and diastolic blood pressure (86 mmHg) than participants with a lower HbA1c (HbA1c <5.9%). Participants in the higher HbA1c were less likely to reside in Coyoacán (the more affluent of the two study districts) than participants with a lower HbA1c (HbA1c <5.9%) (**Table 1**). Among the 110,405 participants aged 35-74 years at recruitment, 28,852 individuals (26%) had ADA-defined prediabetes (HbA1c 5.7%-6.4%) and 7,203 (7%) had IEC-defined prediabetes (HbA1c 6.0%-6.4%). The prevalence of prediabetes (regardless of definition) increased with age (at least up to age 70) and was higher in women than men (at least after age 40) (**Figure 1**).

**Figure 1.**
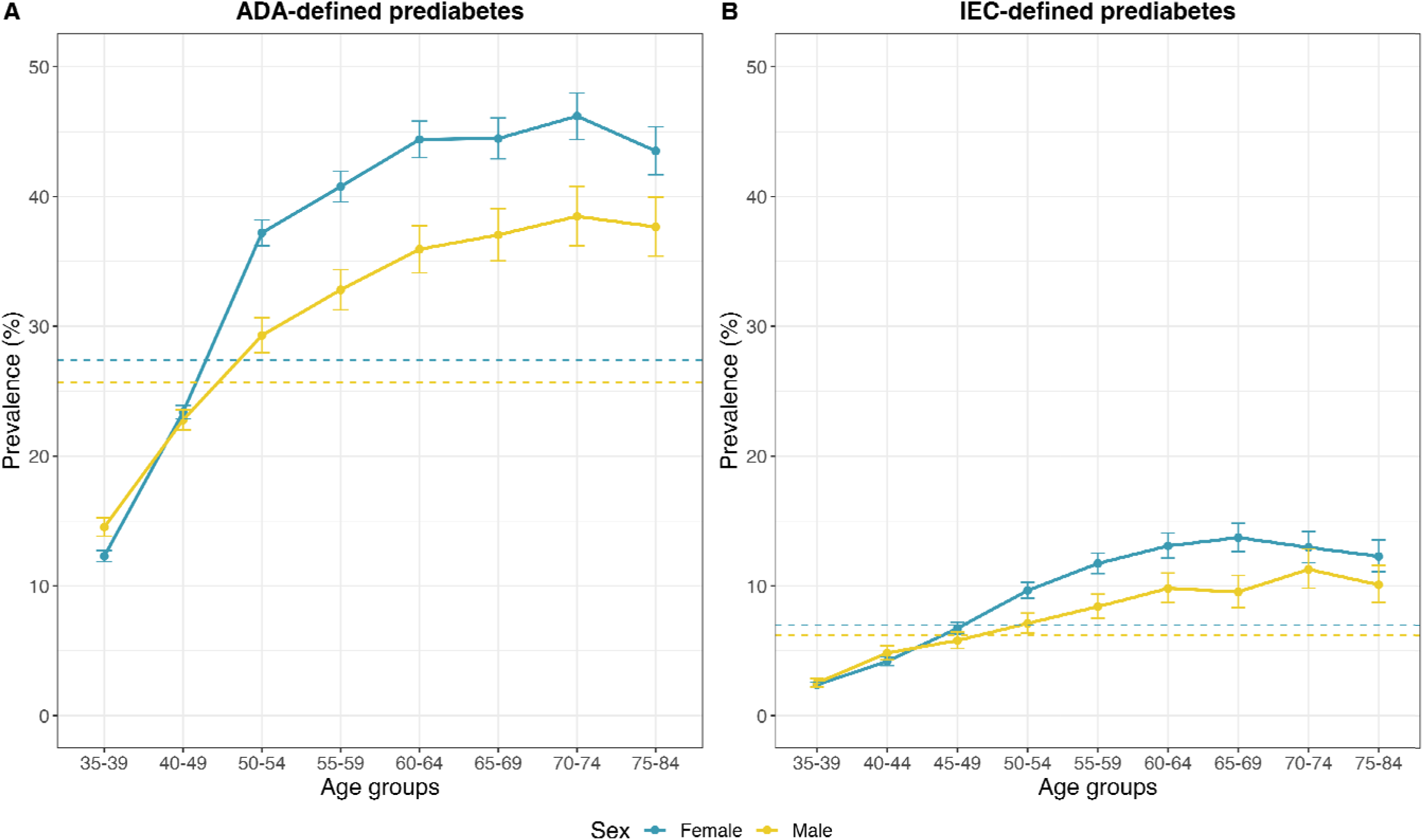
Prevalence of ADA- and IEC-defined prediabetes in 115,919 MCPS participants without previously diagnosed or undiagnosed diabetes, by age and sex. Dashed lines indicate overall prevalence of prediabetes for each definition stratified by sex. ADA: American Diabetes Association. IEC: International Expert Committee. Analyses limited to 115,919 participants aged 35-84, and without previously diagnosed or undiagnosed diabetes (or other chronic disease) at recruitment.

**Table 1.** Baseline characteristics of 110,405 men and women aged 35-74 without previously diagnosed or undiagnosed diabetes or other chronic disease at recruitment.

|  | All participants<br>N = 110,405 | HbA1c, % |  |  |
| --- | --- | --- | --- | --- |
|  |  | <5.7<br>N = 81,553 | 5.7-5.9<br>N = 21,649 | 6.0-6.4<br>N = 7,203 |
| Age (Years) | 49 (10) | 47 (10) | 52 (11) | 54 (10) |
| HbA1c (%) | 5.45 (0.37) | 5.29 (0.26) | 5.83 (0.10) | 6.22 (0.12) |
| Female sex (%) | 74,768 (68%) | 54,829 (67%) | 14,872 (69%) | 5,067 (70%) |
| Residence in Coyoacán (%) | 44,911 (41%) | 37,050 (45%) | 5,986 (28%) | 1,875 (26%) |
| University/college educated (%) | 19,564 (18%) | 16,400 (20%) | 2,486 (11%) | 678 (9.4%) |
| Smoking (%) |  |  |  |  |
| Never | 53,106 (48%) | 38,402 (47%) | 10,910 (50%) | 3,794 (53%) |
| Former | 19,969 (18%) | 14,625 (18%) | 4,033 (19%) | 1,311 (18%) |
| Current | 37,330 (34%) | 28,526 (35%) | 6,706 (31%) | 2,098 (29%) |
| Alcohol intake (%) |  |  |  |  |
| Never | 20,533 (19%) | 14,633 (18%) | 4,391 (20%) | 1,509 (21%) |
| Former | 6,955 (6.3%) | 5,281 (6.5%) | 1,265 (5.8%) | 409 (5.7%) |
| Current | 82,917 (75%) | 61,639 (76%) | 15,993 (74%) | 5,285 (73%) |
| Regular leisure-time physical activity (%) | 25,273 (23%) | 19,812 (24%) | 4,193 (19%) | 1,268 (18%) |
| Body Mass Index (kg/m<sup>2</sup>) | 29.0 (4.9) | 28.3 (4.7) | 30.5 (5.1) | 32.0 (5.3) |
| Waist-to-Height Ratio | 0.60 (0.08) | 0.59 (0.07) | 0.63 (0.08) | 0.66 (0.08) |
| Waist-to-Hip Ratio | 0.89 (0.08) | 0.89 (0.07) | 0.91 (0.07) | 0.92 (0.07) |
| Systolic Blood Pressure (mmHg) | 126 (16) | 125 (16) | 129 (17) | 132 (17) |
| Diastolic Blood Pressure (mmHg) | 83 (11) | 82 (11) | 84 (11) | 86 (11) |

During a median of 18.6 (IQR 17.7-19.8) years’ follow-up, there were 6,810 deaths at ages 35-74 years (including 1,742 from cardiovascular causes, 892 from renal causes and 108 from acute diabetic crises) and 7,067 deaths at ages 75-84 years. All-cause and cause-specific death rates (uniformly age- and sex-standardized) increased according to HbA1c category, with the highest death rates being observed in participants with HbA1c 6.0-6.4% (**Figure 2A**). Given that prediabetes prevalence markedly increased with age, we explored the influence of baseline age and HbA1c categories. For participants aged <50 years at recruitment, HbA1c 6.0%-6.4% was strongly associated with mortality risk, and this association was preserved for participants aged 50-64 years at recruitment; however, we identified that the graded increase in relative risk conferred by higher HbA1c levels was lost in older participants (**Figure 2B)**. When stratified by sex, we observed broadly similar death rate ratios in men and women (**Supplementary** Figure 2). Similar results were obtained when examining deaths occurring at ages 75-84 years (**Supplementary** Figures 2 and 3).

**Figure 2.**
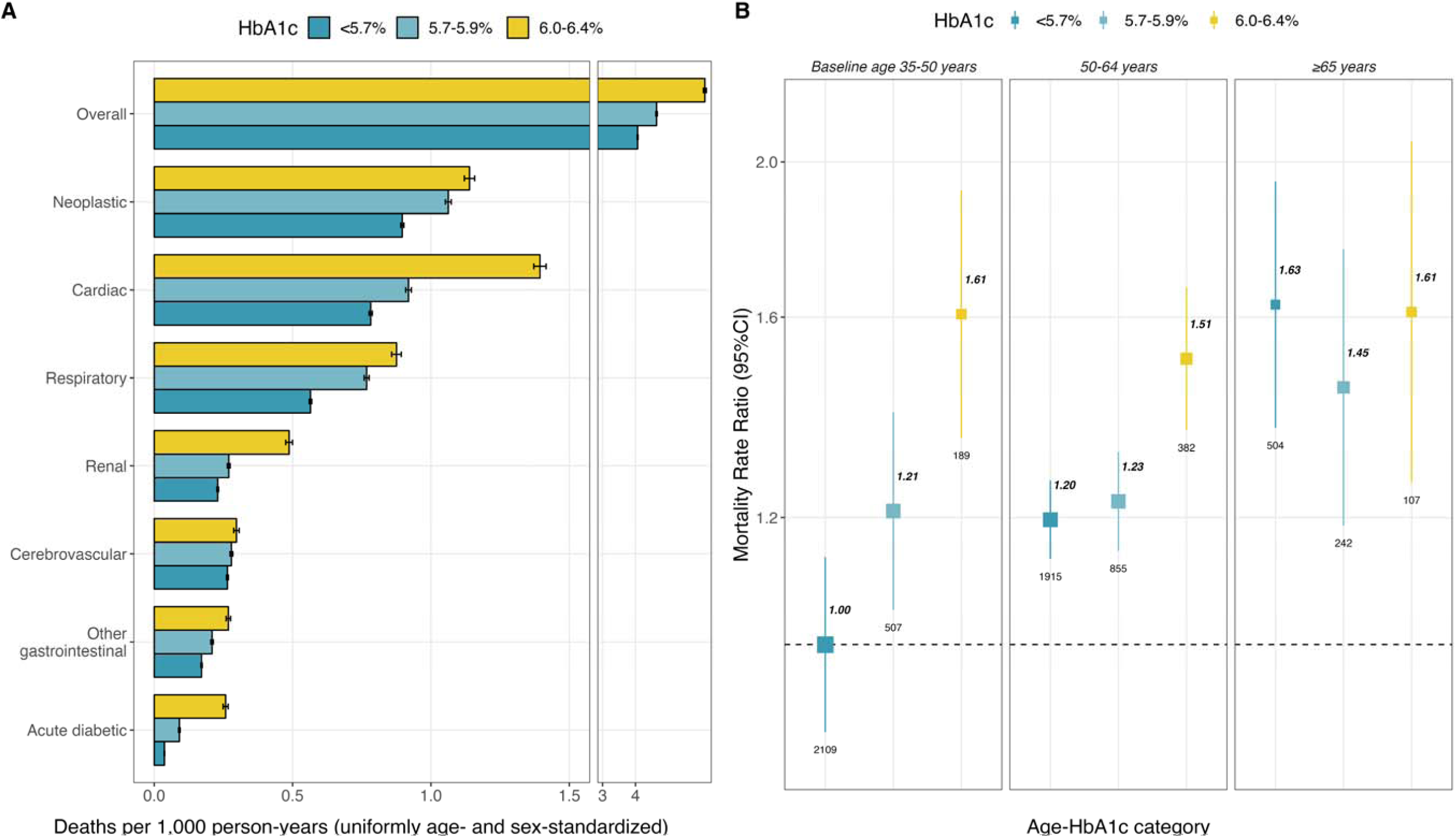
All-cause and cause-specific mortality at ages 35 to 74 years associated with pre-diabetes. Analyses limited to 110,405 participants aged 35-74 and without previously diagnosed or undiagnosed diabetes (or other chronic disease) at recruitment. (A) Uniformly age- and sex-standardized mortality rates per 1,000 person-years according to HbA1c categories, error bars represent 95% confidence intervals. (B) Rate ratios with 95% confidence intervals for all-cause mortality associated with HbA1c categories, stratified by age at recruitment (<50 years, 50-64 years, or ≥65 years). Labels represent the mortality rate ratio (bold) and number of events (plain) for each category, the size of the squares is proportional to the amount of statistical information. Models are stratified by sex and age at risk and adjusted for municipality, education level, physical activity, smoking, and alcohol intake. Each 95% confidence interval reflects the variance of the log risk in that 1 group.

After adjustment for age, sex and confounders, each one percent higher HbA1c was associated with 11% higher risk of death from any cause at ages 35-74 years (RR 1.11 [95% CI 1.03-1.20]) (**Supplementary Table 1**, which also shows the RRs with more and with less adjustment). Compared with individuals without prediabetes, those with ADA-defined prediabetes had 13% higher risk of death at ages 35-74 years (RR 1.13 [95% CI 1.07-1.19]) while those with IEC-defined prediabetes had 28% higher risk (RR 1.28 [95% CI 1.18-1.39]) (**Table 2**). After further adjustment for adiposity these RRs were reduced to 1.06 (1.00-1.13) and 1.16 (1.05-1.27) respectively (**Supplementary Table 1**). Estimates were similar when including participants with other comorbidities at recruitment (**Supplementary Table 2**).

**Table 2:** Excess cause-specific mortality at ages 35-74 years associated with ADA and IEC definitions of prediabetes at recruitment.

| Cause | ADA definition |  |  |  | IEC definition |  |  |  |
| --- | --- | --- | --- | --- | --- | --- | --- | --- |
|  | No. deaths |  | Death RR*<br>(95% CI) | Attributable<br>mortality<br>(%) | No. deaths |  | Death RR* (95%<br>CI) | Attributable<br>mortality<br>(%) |
|  | No<br>prediabetes | Prediabetes |  |  | No<br>prediabetes | Prediabetes |  |  |
| All causes | 4,528 | 2,282 | 1.13 (1.07-1.19) | 4 | 6,132 | 678 | 1.28 (1.18-1.39) | 2 |
| Cardiac | 789 | 452 | 1.24 (1.10-1.41) | 7 | 1,093 | 148 | 1.52 (1.28-1.82) | 4 |
| Cerebrovascular | 257 | 138 | 1.08 (0.87-1.35) | 3 | 362 | 33 | 0.95 (0.66-1.36) | 0 |
| Other vascular | 85 | 61 | 1.49 (1.05-2.13) | 14 | 122 | 24 | 2.10 (1.33-3.32) | 9 |
| All cardiovascular | 1,131 | 651 | 1.22 (1.10-1.35) | 6 | 1,577 | 205 | 1.42 (1.22-1.65) | 3 |
| Renal | 245 | 150 | 1.35 (1.08-1.68) | 10 | 344 | 51 | 1.69 (1.24-2.31) | 5 |
| Acute diabetic crisis | 54 | 54 | 2.63 (1.76-3.94) | 31 | 86 | 22 | 3.43 (2.09-5.62) | 14 |
| Gastrointestinal | 760 | 292 | 0.84 (0.73-0.97) | -5 | 943 | 109 | 1.34 (1.09-1.63) | 3 |
| Neoplastic | 1,063 | 516 | 1.09 (0.97-1.21) | 3 | 1,443 | 136 | 1.09 (0.91-1.30) | 1 |
| Respiratory | 701 | 368 | 1.24 (1.09-1.42) | 7 | 979 | 90 | 1.12 (0.89-1.39) | 1 |
| External/ill-defined/other | 574 | 251 | 1.06 (0.91-1.25) | 2 | 760 | 65 | 1.07 (0.83-1.38) | 1 |

RR=Mortality Rate Ratio; CI=Confidence Interval; ADA definition= HbA1c 5.7-6.4%; IEC definition= HbA1c 6.0-6.4%\*Mortality rate ratio estimates for those with vs without prediabetes at recruitment were stratified by sex and age at risk and adjusted for municipality, education level, physical activity, smoking, and alcohol intake. Analyses excluded data from participants with previously diagnosed or undiagnosed diabetes or other prior chronic disease. \*\*For each definition of prediabetes, the number of deaths attributable to the excess risk associated with it was calculated as $D \times (RR - 1)/RR$ , where D is the proportion of deaths in the prediabetes group and RR is the cause-specific mortality rate ratio for prediabetes versus no prediabetes.

Figure 3 and **Table 2** show the distribution of the 6,810 deaths at ages 35-74 years in participants with and without prediabetes (ADA and IEC definitions), as well as the cause-specific mortality RRs and their 95% CIs. Both ADA- and IEC-defined prediabetes were significantly associated with increased risk of cardiac death, all cardiovascular death, renal death, and death from an acute diabetic crisis. In addition, ADA-defined prediabetes was significantly associated with death from respiratory diseases. When examining deaths occurring at ages 75-84 years, cause-specific RRs tended to be slightly weaker, although both ADA- and IEC-defined prediabetes remained strongly predictive of both renal death and death from acute diabetic crises at these older ages (**Supplementary** Figure 4 and **Supplementary Table 3**).

**Figure 3.**
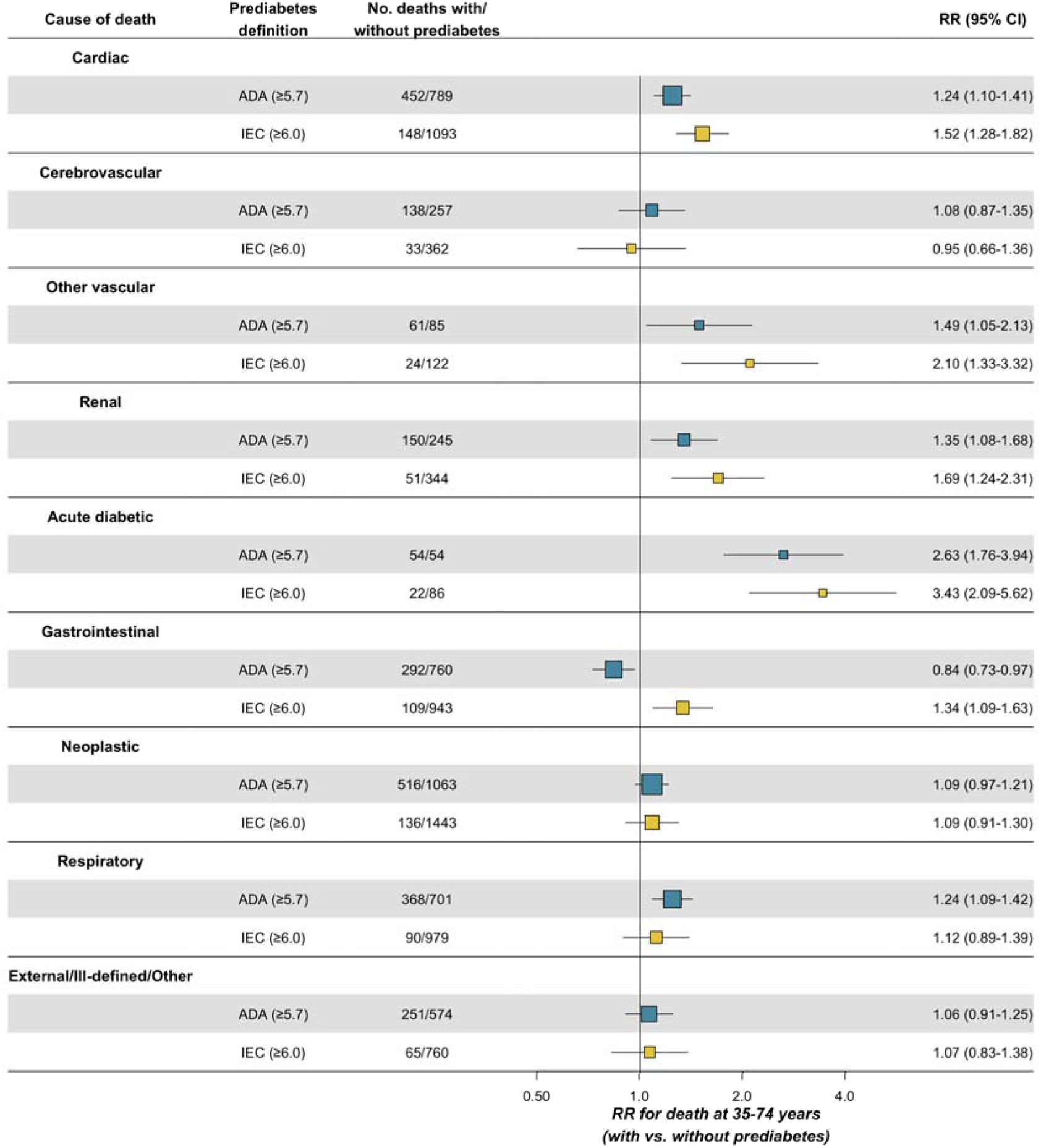
Cause-specific mortality rate ratios at ages 35-74 years associated with ADA- and IEC-defined prediabetes. ADA: American Diabetes Association. IEC: International Expert Committee. RR: Mortality rate ratio. CI: Confidence Interval. Analyses limited to 110,405 participants aged 35-74 and without previously diagnosed or undiagnosed diabetes (or other chronic disease) at recruitment. RRs are stratified by sex and age at risk and adjusted for municipality, education level, physical activity, smoking, and alcohol intake.

Overall, between 35 and 74 years of age in the population without previously diagnosed or undiagnosed diabetes, the excess mortality risk associated with ADA-defined prediabetes accounted for 4% of all deaths, including 6% of cardiovascular deaths, 10% of renal cause deaths, and 31% of deaths from an acute diabetic crisis (**Table 2**). By contrast, the excess mortality risk associated with IEC-defined prediabetes accounted for 2% of all deaths, including 3% of cardiovascular deaths, 5% of renal deaths and 14% of deaths from an acute diabetic crisis.

## DISCUSSION

In this large study of Mexican adults, we explored the association of HbA1c-defined prediabetes and all-cause and cause-specific mortality. Overall, both ADA- and IEC-defined prediabetes were associated with an increased risk of all-cause mortality. The RRs associated with IEC-defined prediabetes (HbA1c 6%-6.4%) were somewhat larger than those associated with ADA-defined prediabetes (HbA1c 5.7%-6.4%). When considering cause-specific mortality, both definitions were associated with an increased risk of mortality from cardiovascular disease, kidney disease and acute diabetic crises, but only the ADA definition was significantly associated with respiratory-related mortality, perhaps reflecting increased statistical power for this definition. These results support the view that prediabetes is a significant risk factor for cardiometabolic mortality.

A recently updated meta-analysis of prospective cohorts reported a 13% increase in the risk of all-cause mortality when considering any definition of prediabetes^3^. When examining definitions based on HbA1c, the IEC definition yielded a 21% increase in risk, while the ADA definition was not significantly associated with mortality^3^. In the present study, we found that prediabetes was significantly associated with increased risk of premature death from cardiovascular disease, kidney disease and acute diabetic crises irrespective of the definition used. This is in line with previous reports from other populations although, unlike previous reports, we did not find any significant association with cancer death.^3,26–28^ Notably, estimates of mortality associated with ADA and IEC definitions were reasonably consistent, with differences in attributable mortality being a function of prevalence more than associated mortality risks. Expanding prediabetes screening has been challenging given that preventive efforts should not only be aimed at predicting progression to type 2 diabetes, but also risk of all-cause and cause-specific mortality^2,3,20^. Our findings support the view that the IEC definition of prediabetes could be more useful at identifying individuals at the highest risk for cause-specific mortality compared to the ADA definition, at the same time limiting potential adverse impacts on individuals and healthcare systems of labelling a large proportion of the population with a diagnosis of prediabetes^1^. However, selecting the most optimal point for screening should also balance the relevance of additional outcomes, perhaps most importantly type 2 diabetes.

The significance of prediabetes as a risk factor for all-cause and cause-specific mortality lies mainly within its high prevalence worldwide^29^. Previous efforts from our group reported an increasing prevalence of HbA1c-defined prediabetes in Mexico between 2016 and 2022^7^. Notably, the predominantly urban MCPS sample in the current report yielded a high prevalence of prediabetes (one quarter of those without diabetes had ADA-defined prediabetes), similar to that estimated in a previous cross-sectional study of adults in Mexico City (26% of the adult population as a whole)^30^. In the present analysis, we observed that among those without diabetes 6% of the premature cardiovascular deaths, 10% of the premature renal deaths and 31% of the premature acute diabetic deaths could be attributed to the long-term hazards associated with ADA-defined prediabetes. Interestingly, we also identified that the magnitude of the risk ratio of prediabetes for all-cause and cause-specific mortality decreased at older ages, being consistent with findings from the ARIC study, which identified that regression to normoglycemia from prediabetes was more common in this population, potentially contributing to the decrease in at-risk population over time^31^.

Our study has several strengths, including its large sample size, prospective design, and prolonged follow-up, which allowed us to evaluate long-term prediabetes-related outcomes compared with previous studies. However, we also acknowledge some limitations which should be considered to adequately frame our results. First, by only using HbA1c-based definitions of prediabetes, we were unable to assess separately the mortality risks associated with impaired fasting glucose and/or glucose intolerance; this may limit our ability to identify differential risk of adverse outcomes based on the specific definition employed^4,5,32^. Nevertheless, by exploring both ADA and IEC HbA1c-based definitions we contribute valuable insights to inform debate regarding the most appropriate definition of prediabetes for predicting future cardiometabolic disease risks. Prediabetes has also been recognized to increase the risk of a cluster of multiple chronic comorbidities, which may underlie the associations observed for mortality^33^. To isolate the effect of prediabetes, we excluded individuals with comorbidities from our main analyses and focused on premature death to reduce the potential influence of aging in our estimations. We expect that these adjustments were more likely to generate results which were causal, but we cannot rule out the possibility of residual confounding or reverse causality in our findings. Furthermore, due to study design, we did not have prospective data to identify participants with prediabetes who developed diabetes or other non-fatal events during follow-up; this prevented us from analyzing the relevance of prediabetes for mortality and cardiometabolic risks independent of its association with increased risk to progression to diabetes, an area which will require further evaluations with longitudinal assessments. HbA1c levels have been known to be impacted over time by individual-level factors, including chronic diseases, kidney, and liver function^34^; this effect may have been mitigated by excluding individuals with comorbidities, but there remains a possibility of unconsidered factors influencing HbA1c levels over time. Lastly, we acknowledge that MCPS data primarily arises from two districts in Mexico City and our estimates may not be representative of the whole Mexican population; however, large-scale prospective studies of non-representative cohorts of individuals such as MCPS can provide reliable evidence about the associations of risk factors with disease that are widely generalizable^35,36^.

In this Mexican population prediabetes was significantly associated with increased risks of cardiovascular, renal, and acute diabetic mortality. Given the increased risks associated with prediabetes, coupled with its high prevalence in Mexican adults, population-wide screening strategies to facilitate its timely diagnosis, alongside implementation of strategies to prevent and delay associated cardiometabolic diseases and related mortality, should be considered. However, further work is needed to identify the appropriate outcome-driven definitions of prediabetes to promote efficient screening and prevention strategies that minimize overdiagnosis and improve cardiometabolic outcomes.

## ACKNOWLEDGMENTS

This project was registered and approved by the Research Committee at Instituto Nacional de Geriatría, project number DI-PI-006/2020. CAFM is enrolled at the PECEM Program of the Faculty of Medicine at UNAM. CAFM and DRG are supported by CONAHCYT. JAS was supported by Grant Number K23DK135798 from the NIH/NIDDK and by the Massachusetts General Hospital Executive Committee and Center for Diversity and Inclusion Physician-Scientist Development Award. The authors thank the participants for their willingness to take part in this prospective study 20 years ago. This research was conducted using Mexico City Prospective Study (MCPS) data obtained through an open-access data request (application number 2022-012).

## AUTHOR CONTRIBUTIONS

Establishing the cohort: PKM, JAD and RTC. Obtaining funding: JBC, JRE, PKM, JAD, RTC and OYBC. Data acquisition, analysis, or interpretation of data: CAFM, OYBC, CDPC, JPE, DRG, LFC, AVV, PSC, ANL, MRBA, JPDS, JBC, JRE, PKM, RTC, JAD, JAS, NEAV. Drafting first version of manuscript: CAFM, OYBC. Critical revision of the report for important intellectual content: All authors. All authors have seen and approved the final version and agreed to its publication. Each author contributed important intellectual content during manuscript drafting or revision and accepts accountability for the overall work by ensuring that questions pertaining to the accuracy or integrity of any portion of the work are appropriately investigated and resolved.

## DATA AVAILABILITY

Data from the Mexico City Prospective Study are available to bona fide researchers. For more details, the study’s Data and Sample Sharing policy may be downloaded (in English or Spanish) from https://www.ctsu.ox.ac.uk/research/mcps. Available study data can be examined in detail through the study’s Data Showcase, available at https://datashare.ndph.ox.ac.uk/mexico/. Code is available for reproducibility of results at https://github.com/oyaxbell/prediabetes_mcps/.

## RIGHTS RETENTION STATEMENT

For the purposes of open access, the authors have applied a Creative Commons Attribution (CC BY) licence to any Author Accepted Manuscript version arising.

