## Supplementary Material for "Prediabetes as a risk factor for all-cause and cause-specific mortality: a prospective analysis of 115,919 adults without diabetes in Mexico City"

**SUPPLEMENTARY METHODS**

*ICD-10 codes for cause-specific mortality, indicating number of deaths in brackets.*

| ***Cause-specific mortality, Number of deaths*** | ***ICD-10 codes*** |
| --- | --- |
| *Cardiac deaths, N=3297* | *I018(1), I050(4), I051(1), I059(16), I070(1), I071(3), I080(5), I081(1), I091(1), I099(14), I110(163)I119(29), I200(2), I209(6), I210(4), I211(10), I213(1), I219(2430)I220(1), I221(2), I229(1), I248(1)I249(19), I251(58), I255(4), I258(7), I259(112), I270(8), I272(3), I279(12), I289(1), I301(1), I318(1), I330(4), I340(5), I348(1), I350(13), I351(1), I358(3), I359(1), I38X(9), I420(15), I429(1), I441(1)I442(5), I443(2), I459(1), I469(6), I471(2), I472(1), I480(1), I489(8), I48X(13), I490(8), I499(16), I500(73), I501(12), I509(119), I518(3), I519(8), Q210(1), Q231(1), Q238(1), Q248(1), R570(38)* |
| *Cerebrovascular deaths, N=990* | *F019(10), I600(2), I602(1), I608(1), I609(81), I610(1), I613(1), I614(1), I615(1), I618(1), I619(234), I620(11), I629(4), I633(4), I634(27), I635(13), I638(1), I639(97), I64X(136), I652(1), I671(11), I672(1), I673(1), I674(4), I678(132)I679(146)I690(2), I691(1), I693(10), I694(16), I698(38)* |
| *Other vascular deaths, N=294* | *E115(4), E145(4), I260(4), I269(87), I709(2), I710(3), I712(1), I713(8), I714(9), I718(1), I719(2), I729(1), I739(5), I740(1), I741(1), I771(12), I776(1), I779(1), I802(6), I803(2), I822(1), I828(2), I829(4), I872(8), I879(1), I888(1), I890(1), I99X(1), K550(98), K552(1), K559(20, )K761(1)* |
| *Renal deaths, N=892* | *E102(2), E112(110), E142(37), I120(165), I129(3), I130(2), I131(1), I132(17), N002(1), N009(6), N039(11), N059(9), N10X(1), N12X(3), N133(1), N151(9), N170(2), N179(105), N180(9), N185(7), N189(156), N19X(28), N200(5), N201(1), N202(1), N289(1), N300(1), N308(1), N309(3), N390(190), Q612(1), Q613(2), Q619(1)* |
| *Acute diabetic deaths, N=186* | *E110(61), E111(59), E140(34), E141(26), E162(6)* |
| *Hepatobiliary deaths, N=1033* | *B169(2), B171(20), B181(2), B182(13), B189(1), B190(2), B199(2), D134(1), D136(1), I850(12), I859(4), K701(20), K702(1), K703(151), K704(22), K709(14), K711(1), K716(1), K720(9), K721(70), K729(195, )K740(1), K742(1), K743(3), K745(3), K746(271), K750(11), K754(2), K759(1), K760(3), K764(1), K766(11), K767(12), K768(1), K769(14), K800(7), K801(7), K802(2), K803(6), K805(1), K810(18), K811(10), K819(6), K820(1), K821(1), K829(5), K830(22), K831(3), K851(3), K852(3), K858(1), K859(36), K85X(18), K861(2), K863(1), Q447(1)* |
| *Other gastrointestinal deaths, N=766* | *A047(3), A090(26), A099(47), A09X(9), A183(1), K052(1), K088(1), K102(1), K137(1), K20X(2), K219(1), K223(3), K228(1), K251(5), K254(22), K255(13), K256(2), K259(11), K264(12), K265(7), K266(1), K269(5), K272(1), K274(2), K275(3), K290(11), K291(3), K292(1), K295(8), K296(1), K297(7), K311(2), K318(11), K319(1), K352(5), K353(4), K358(2), K37X(4), K389(1), K403(7), K404(2), K409(2), K413(3), K419(1), K420(6), K421(1), K429(2), K430(2), K431(1), K439(3), K460(6), K461(1), K469(3), K509(1), K513(1), K519(2), K529(4), K560(3), K562(5), K566(78), K567(2), K572(3), K573(4), K578(13), K579(22), K593(8), K610(3), K611(1), K628(1), K631(23), K632(5), K638(2), K639(7), K650(28), K658(1), K659(54), K914(1), K918(1), K920(47), K921(4), K922(159), K931(1)* |
| *Neoplastic deaths, N=2428* | *C009(1), C01X(1), C029(12), C049(1), C059(1), C069(4), C07X(5), C089(1), C099(1), C109(4), C119(2), C139(1), C140(7), C159(21), C160(4), C169(216)C170(14), C179(2), C181(1), C182(1), C183(1), C187(3), C189(132), C19X(8), C20X(20), C210(1), C220(53), C221(24), C227(1), C229(112), C23X(33), C240(9), C241(7), C248(3), C249(18), C250(25), C258(1), C259(117)C260(1), C269(2), C310(1), C311(1), C319(1), C329(15), C33X(2), C340(1), C349(185)C37X(1), C382(1), C383(2), C384(1), C402(1), C410(1), C412(2), C414(1), C419(7), C435(1), C437(2), C439(21), C442(1), C443(1), C444(3), C445(1), C447(1), C449(10), C450(2), C451(2), C457(1), C459(4), C469(1), C479(1), C480(11)C482(6), C490(1), C492(2), C493(1), C495(1), C499(14), C509(183), C519(5), C52X(1), C530(1), C539(111), C541(25), C549(2), C55X(12), C56X(102), C609(2), C61X(157), C629(3), C64X(89), C65X(1), C679(28), C680(2), C689(1), C709(1), C710(17), C711(1), C718(3), C719(41), C720(1), C729(1), C73X(30), C749(1), C751(1), C753(1), C759(2), C760(13), C762(7), C763(3), C764(1), C767(1), C780(6), C786(2), C787(14), C788(3), C790(1), C793(3), C794(2), C795(1)C796(1), C798(4), C800(36), C809(21), C80X(5), C811(1), C817(1), C819(11), C833(15), C839(2), C844(1), C845(3), C851(1), C857(1), C859(57), C880(1), C900(53), C902(1), C910(25), C911(3), C919(2), C920(32), C921(5), C924(1), C927(4), C929(5), C930(1), C950(1), C959(5), C969(1), C97X(2), D371(3), D372(1), D374(2), D376(8), D377(7)D380(1), D381(6), D383(2), D391(3), D397(1), D410(1), D414(1), D430(12), D431(2), D432(1), D449(1), D483(1), D484(1), D486(1), D487(6), D489(1)* |
| *Respiratory deaths, N=2450* | *A150(1), A162(11), B206(2), B909(1), E840(1), E848(1), J069(1), J09(2), J100(3), J129(6), J150(1), J151(2), J152(2), J157(1), J159(41), J180(84), J181(49), J182(2), J189(706), J209(12), J22X(24), J348(1), J399(1), J40X(2), J42X(29), J439(68), J440(245)J441(8), J448(13), J449(369), J459(20), J46X(2), J47X(3), J64X(10), J65X(1), J679(2), J680(1), J684(1), J690(2), J80X(5), J81X(8), J841(104), J849(10), J852(1), J869(2), J90X(5), J920(1), J939(1), J960(7), J961(3), J969(3), J980(1), J981(1), J984(26), J985(2), J988(14), J989(2), Q311(1), U071(266), U072(256)* |
| *Ill-defined or external, N=1541* | *A181(1), A182(1), A199(3), A415(1), A418(1), A419(147), A810(2), A86X(1), B200(1), B201(1), B207(3), B208(6), B210(1), B212(1), B218(1), B227(2), B238(4), B24X(4), B465(2), B690(2), B699(1), B948(1), B99X(1), D033(1), D181(1), D27X(1), D329(9), D464(1), D469(11), D479(1), D529(1), D530(1), D594(1), D619(4), D62X(1), D649(9), D65X(3), D682(1), D691(1), D693(2), D694(1), D696(2), D699(3), D762(1), E031(1), E035(1), E039(10), E049(1), E055(1), E059(3), E116(1), E119(4), E149(3), E230(1), E249(2), E279(1), E43X(5), E440(1), E46X(7), E500(1), E509(1), E660(1), E835(1), E86X(7), E870(1), E871(1), E872(12), E875(2), E878(8), E889(2), F03X(26), F09X(1), F102(6), F182(1), F209(1), F329(3), G009(2), G039(2), G049(6), G060(2), G10X(6), G121(1), G122(14), G20X(29)G219(1), G301(7), G309(16), G310(1), G312(1), G319(1), G35X(3), G379(1), G403(2), G406(1), G409(12), G419(2), G439(1), G473(1), G589(2), G603(1), G610(4), G700(1), G709(1), G710(1), G809(1), G822(1), G919(5), G931(8), G934(7), G935(1), G936(1), G937(1), G939(1), G958(2), G959(1), I10X(4), L020(1), L021(2), L022(1), L023(2), L031(3), L032(1), L039(5), L088(1), L089(31), L100(2), L108(1), L109(1), L512(5), L899(20), L89X(1), L905(6), L921(1), L984(14), L988(1), L989(1), M009(3), M050(2), M068(1), M069(25), M109(1), M139(1), M165(1), M199(1), M300(1), M311(1), M313(1), M319(1), M321(4), M329(3), M331(2), M349(1), M350(1), M471(1), M623(20), M726(3), M798(25), M809(1), M819(3), M869(1), M993(1), N40X(16), N410(1), N498(4), N499(1), N719(1), N739(3), N823(1), N939(1), N948(1), O720(1), R048(1), R092(2), R100(2), R13X(1), R190(1), R54X(1), R571(12), R578(2), R579(2), R58X(6), R64X(2), R688(21), R69X(1), R91X(1), R99X(307)S720(1), S729(2), T07X(1), V011(1), V031(1), V049(1), V051(1), V093(3), V099(89), V149(1), V209(1), V299(1), V439(1), V494(1), V499(9), V580(1), V581(1), V719(1), V785(1), V878(5), V892(6), V899(5), W018(1), W040(1), W050(1), W100(15), W104(2), W105(1), W108(2), W126(1), W130(14), W134(3), W135(1), W138(1), W139(2), W170(4), W172(1), W174(2), W176(1), W178(1), W179(1), W180(6), W181(1), W184(3), W188(1), W189(1), W190(15), W194(2), W199(4), W200(1), W206(1), W228(1), W250(1), W314(1), W340(1), W370(1), W557(1), W704(1), W744(1), W748(2), W769(1), W780(3), W789(1), W808(1), W849(2), W87(1), W871(1), W878(1), X09(1), X090(3), X094(1), X219(1), X440(1), X449(1), X459(1), X470(1), X530(1), X590(6), X594(1), X598(1), X599(39), X650(1), X680(1), X700(5), X702(1), X708(1), X740(4), X780(2), X910(2), X912(1), X914(2), X950(4), X954(15), X955(1), X959(1), X990(6), X994(5), X999(2), Y018(1), Y044(2), Y084(1), Y090(2), Y094(1), Y099(1), Y159(1), Y200(1), Y239(1), Y240(1), Y244(3), Y245(1), Y248(1), Y249(1), Y260(2), Y265(1), Y280(1), Y330(1), Y334(2), Y338(1), Y340(3), Y344(9), Y345(1), Y346(1), Y348(3), Y349(18), Y579(2), Y609(1), Y839(3), Y86X(3), Y899(1)* |

**Supplementary Figure 1. Flowchart of study participant selection in individuals included in the baseline 1998-2004 evaluation of the Mexico City Prospective Study.** IHD: Ischemic heart disease. CKD: Chronic kidney disease. COPD: Chronic obstructive pulmonary disease. MCPS: Mexico City Prospective Study.

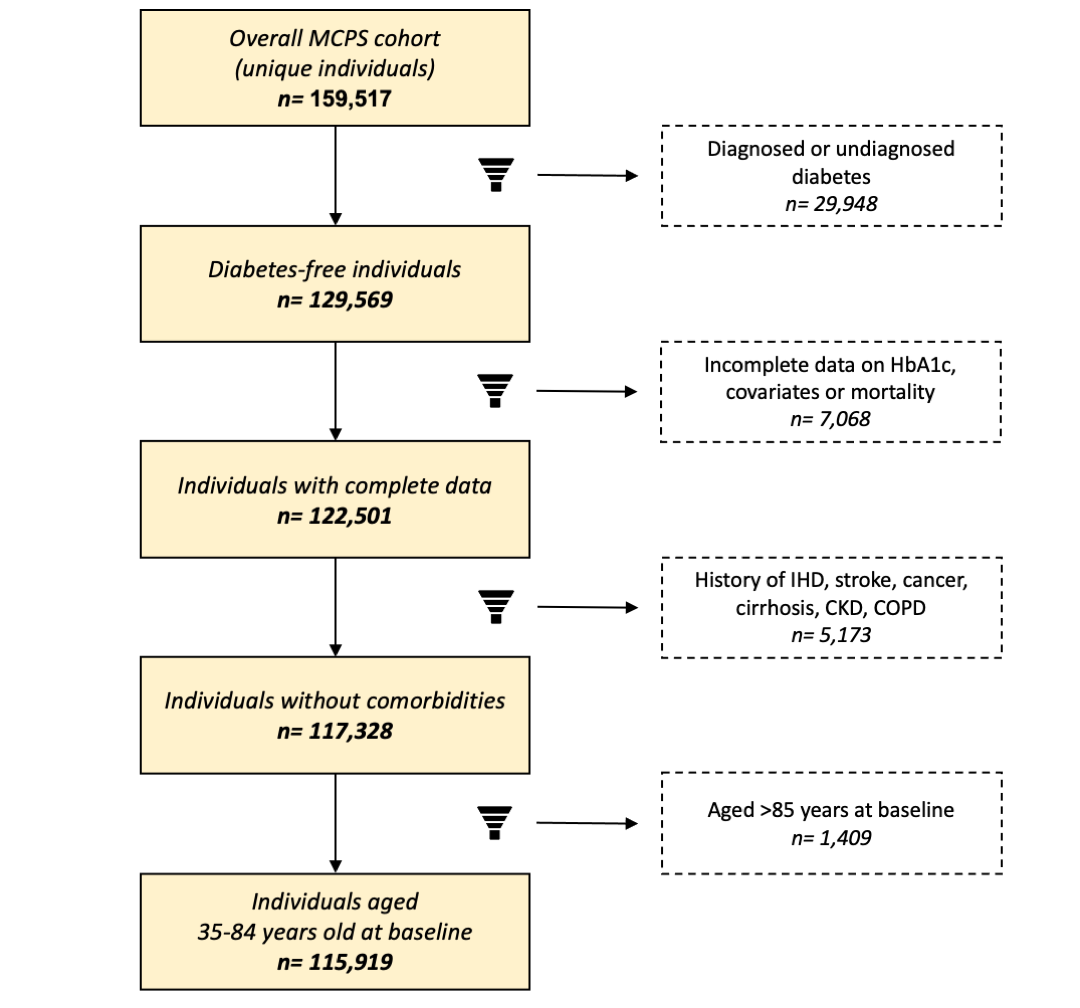

**Supplementary Figure 2. Overall mortality rate ratios at ages 35-74 years and 75-84 years at different levels of HbA1c stratified by sex.** Rate ratios with 95% confidence intervals for all-cause mortality associated with HbA1c categories, stratified by sex. Labels represent the mortality rate ratio (bold) and number of events (plain) for each category, the size of the squares is proportional to the amount of statistical information. Models are stratified by sex and age at risk and adjusted for municipality, education level, physical activity, smoking, and alcohol intake. Each 95% confidence interval reflects the variance of the log risk in that 1 group.

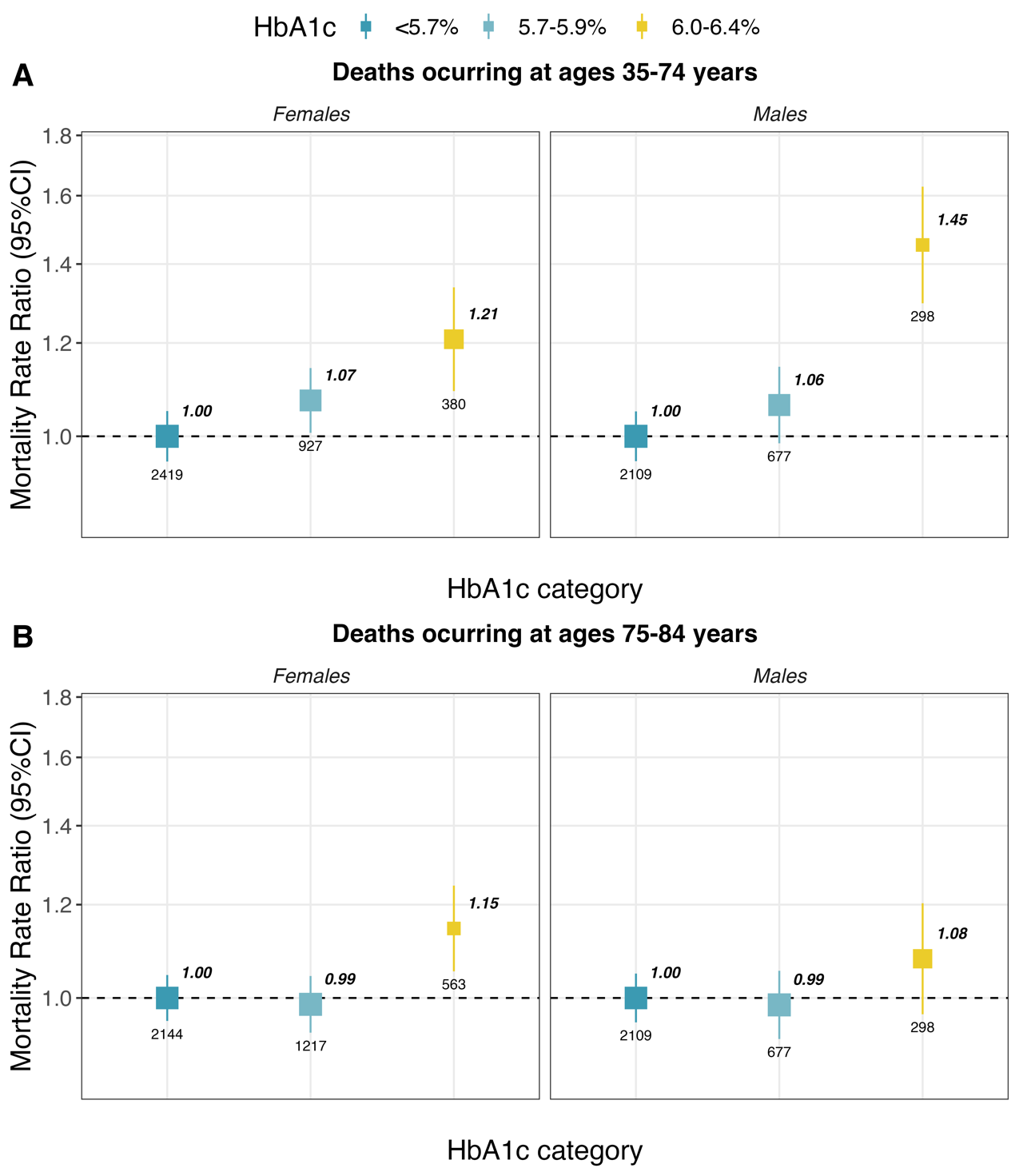

**Supplementary Figure 3. Overall and cause-specific mortality rates and mortality rate ratios at ages 75-84 years and at different levels of HbA1c.** Analyses limited to 7,067 deaths at ages 75-84 among 30,114 participants aged 75-84 years at the end of follow-up who did not have previously diagnosed or undiagnosed diabetes (or other chronic disease) at recruitment. (A) Uniformly age- and sex-standardized mortality rates per 1,000 person-years according to HbA1c categories, error bars represent 95% confidence intervals. (B) Rate ratios with 95% confidence intervals for all-cause mortality associated with HbA1c categories, stratified by age at recruitment (<50 years, 50-64 years, or ≥65 years). Labels represent the mortality rate ratio (bold) and number of events (plain) for each category, the size of the squares is proportional to the amount of statistical information. Models are stratified by sex and age at risk and adjusted for municipality, education level, physical activity, smoking, and alcohol intake. Each 95% confidence interval reflects the variance of the log risk in that 1 group.

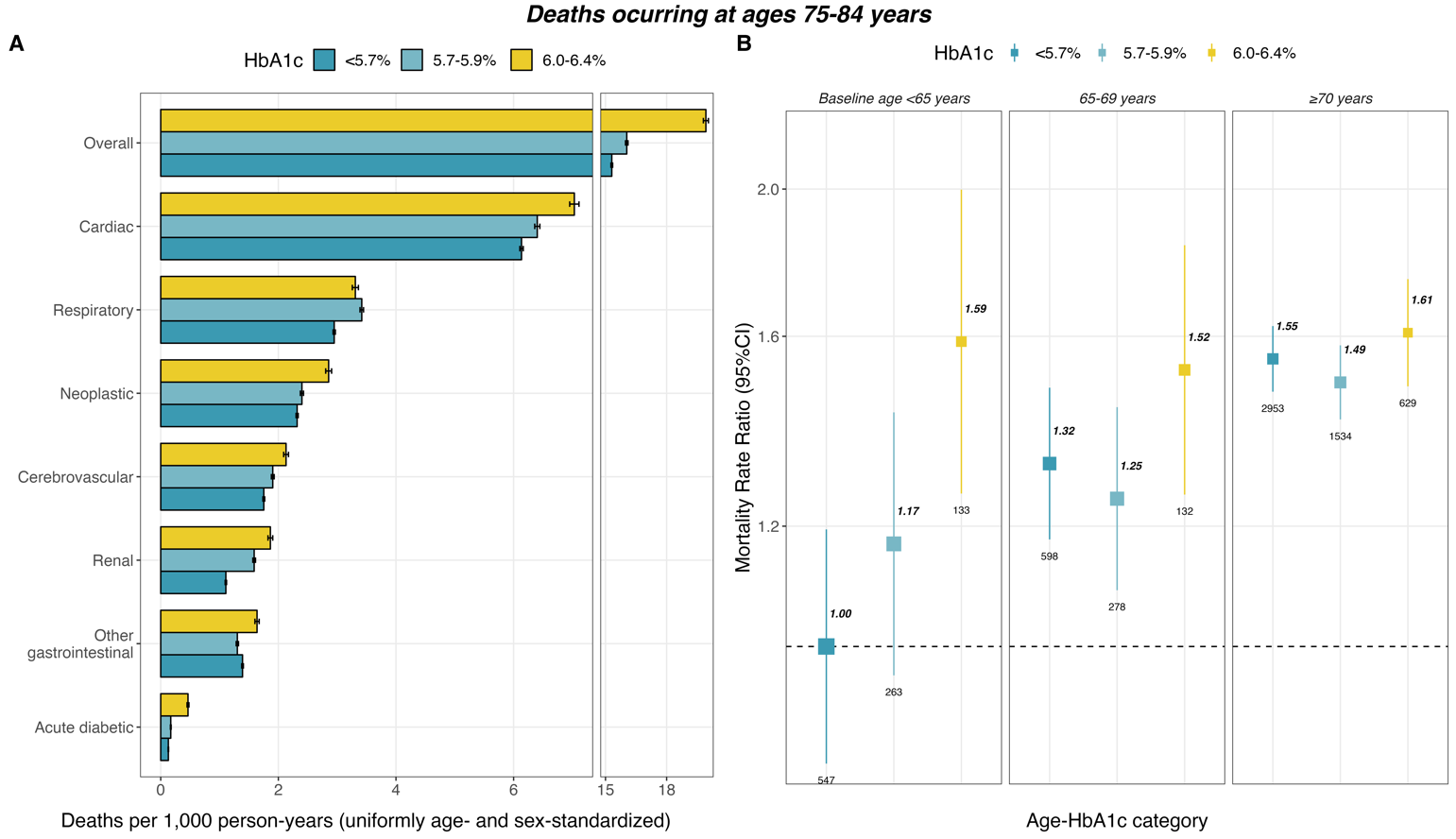

**Supplementary Figure 4.** **Cause-specific mortality rate ratios at ages 75-84 years associated with ADA- and IEC-defined prediabetes.** ADA: American Diabetes Association. IEC: International Expert Committee. RR: Mortality rate ratio. CI: Confidence Interval. RRs are stratified by sex and age at risk and adjusted for municipality, education level, physical activity, smoking, and alcohol intake.

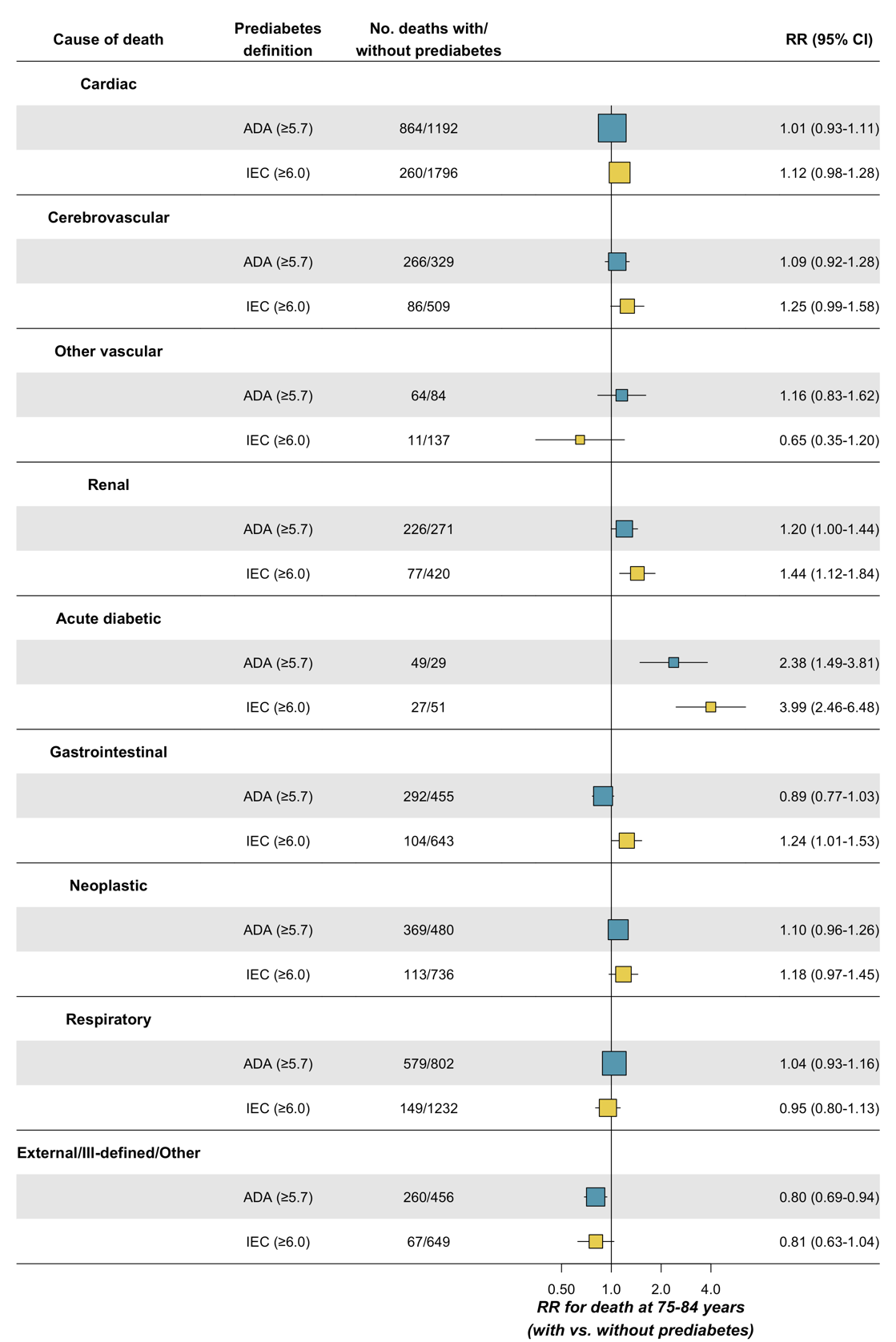

**Supplementary Table 1. Association of HbA1c and ADA- and IEC-defined prediabetes with all-cause mortality at ages 35-74 years.** ADA: American Diabetes Association. IEC: International Expert Committee. RR: Mortality rate ratio. CI: Confidence Interval. BMI: body mass index. WHtR: waist-to-height ratio. Analyses limited to 110,405 participants aged 35-74 and without previously diagnosed or undiagnosed diabetes (or other chronic disease) at recruitment. All models are stratified by sex and age at risk in 5-year increments using the Lexis expansion.

|  | | **Age-at-risk, sex, and district of residence** | **+ Education, physical activity, smoking, and alcohol intake** | **+ BMI and WHtR** |
| --- | --- | --- | --- | --- |
|  | **Predictor** | **RR (95% CI)** | **RR (95% CI)** | **RR (95% CI)** |
| **Continuous**  **(per 1 % unit)** | HbA1c (%) | 1.15 (1.07, 1.24) | 1.11 (1.03, 1.20) | 1.07 (0.98, 1.16) |
| **HbA1c category** | *<5.7%* | 1.00 | 1.00 | 1.00 |
|  | *5.7-5.9%* | 1.09 (1.03, 1.16) | 1.07 (1.01, 1.13) | 1.03 (0.96, 1.10) |
|  | *6.0-6.4%* | 1.34 (1.23, 1.46) | 1.31 (1.20, 1.42) | 1.17 (1.06, 1.29) |
| **Prediabetes definition** | ADA (5.7-6.4%) | 1.15 (1.09, 1.22) | 1.13 (1.07, 1.19) | 1.06 (1.00, 1.13) |
|  | IEC (*6.0-6.4%)* | 1.30 (1.20, 1.41) | 1.28 (1.18, 1.39) | 1.16 (1.05, 1.27) |

**Supplementary Table 2. Association of HbA1c and ADA- and IEC-defined prediabetes with all-cause mortality at 35-74 years including individuals with comorbidities.** ADA: American Diabetes Association. IEC: International Expert Committee. RR: Mortality rate ratio. CI: Confidence Interval. BMI: body mass index. WHtR: waist-to-height ratio. Analyses limited to 114,765 aged 35-74 and without previously diagnosed or undiagnosed diabetes, but with other chronic diseases at baseline. All models are stratified by sex and age at risk in 5-year increments using the Lexis expansion.

|  | | **Age-at-risk, sex, and district of residence** | **+ Education, physical activity, smoking, and alcohol intake** | **+ BMI and WHtR** |
| --- | --- | --- | --- | --- |
|  | **Predictor** | **RR (95% CI)** | **RR (95% CI)** | **RR (95% CI)** |
| **Continuous**  **(per 1 % unit)** | HbA1c (%) | 1.28 (1.18, 1.39) | 1.23 (1.13, 1.34) | 1.04 (0.95, 1.12) |
| **HbA1c category** | *<5.7%* | 1.00 | 1.00 | 1.00 |
|  | *5.7-5.9%* | 1.14 (1.07, 1.22) | 1.12 (1.05, 1.19) | 1.02 (0.95, 1.09) |
|  | *6.0-6.4%* | 1.42 (1.30, 1.56) | 1.39 (1.26, 1.52) | 1.18 (1.07, 1.29) |
| **Prediabetes definition** | ADA (5.7-6.4%) | 1.21 (1.14, 1.28) | 1.18 (1.12, 1.26) | 1.06 (1.00, 1.12) |
|  | IEC (*6.0-6.4%)* | 1.37 (1.25, 1.49) | 1.34 (1.23, 1.46) | 1.17 (1.07, 1.28) |

**Supplementary Table 3. Association of HbA1c and ADA- and IEC-defined prediabetes with all-cause mortality at ages 75-84 years in individuals without comorbidities.** ADA: American Diabetes Association. IEC: International Expert Committee. RR: Mortality rate ratio. CI: Confidence Interval. BMI: body mass index. WHtR: waist-to-height ratio. Analyses limited to 30,114 participants aged 75-84 years at the end of follow-up and without previously diagnosed or undiagnosed diabetes (or other chronic disease) at baseline. All models are stratified by sex and age at risk in 5-year increments using the Lexis expansion.

|  | | **Age-at-risk, sex, and district of residence** | **+ Education, physical activity, smoking, and alcohol intake** | **+ BMI and WHtR** |
| --- | --- | --- | --- | --- |
|  | **Predictor** | **RR (95% CI)** | **RR (95% CI)** | **RR (95% CI)** |
| **Continuous**  **(per 1 % unit)** | HbA1c (%) | 0.99 (0.93, 1.06) | 0.98 (0.92, 1.05) | 0.94 (0.88, 1.00) |
| **HbA1c category** | *<5.7%* | 1.00 | 1.00 | 1.00 |
|  | *5.7-5.9%* | 0.98 (0.93, 1.03) | 0.98 (0.93, 1.03) | 0.95 (0.90, 1.00) |
|  | *6.0-6.4%* | 1.12 (1.04, 1.20) | 1.11 (1.04, 1.20) | 1.07 (0.99, 1.14) |
| **Prediabetes definition** | ADA (5.7-6.4%) | 1.02 (0.97, 1.07) | 1.01 (0.97, 1.06) | 0.98 (0.94, 1.03) |
|  | IEC (*6.0-6.4%)* | 1.13 (1.05, 1.21) | 1.12 (1.05, 1.20) | 1.09 (1.01, 1.16) |
